# Diversity and selection analyses identify transmission-blocking antigens as the optimal vaccine candidates in *Plasmodium falciparum*

**DOI:** 10.1101/2024.05.11.24307175

**Authors:** Ilinca I. Ciubotariu, Bradley K. Broyles, Shaojun Xie, Jyothi Thimmapuram, Mulenga C. Mwenda, Brenda Mambwe, Conceptor Mulube, Japhet Matoba, Jessica L. Schue, William J. Moss, Daniel J. Bridges, He Qixin, Giovanna Carpi

**Author notes:** **Corresponding authors:** Correspondence to Giovanna Carpi and Qixin He. Joint senior authors (He Qixin, Giovanna Carpi).

## Abstract

**Background:** A highly effective vaccine for malaria remains an elusive target, at least in part due to the under-appreciated natural parasite variation. This study aimed to investigate genetic and structural variation, and immune selection of leading malaria vaccine candidates across the *Plasmodium falciparum*’s life cycle.

**Methods:** We analyzed 325 *P. falciparum* whole genome sequences from Zambia, in addition to 791 genomes from five other African countries available in the MalariaGEN Pf3k Rdatabase. Ten vaccine antigens spanning three life-history stages were examined for genetic and structural variations, using population genetics measures, haplotype network analysis, and 3D structure selection analysis.

**Findings:** Among the ten antigens analyzed, only three in the transmission-blocking vaccine category display *P*. *falciparum* 3D7 as the dominant haplotype. The antigens *AMA1, CSP, MSP1_19_* and *CelTOS,* are much more diverse than the other antigens, and their epitope regions are under moderate to strong balancing selection. In contrast, *Rh5*, a blood stage antigen, displays low diversity yet slightly stronger immune selection in the merozoite-blocking epitope region. Except for *CelTOS*, the transmission-blocking antigens *Pfs25*, *Pfs48/45*, *Pfs230*, *Pfs47*, and *Pfs28* exhibit minimal diversity and no immune selection in epitopes that induce strain-transcending antibodies, suggesting potential effectiveness of 3D7-based vaccines in blocking transmission.

**Interpretations:** These findings offer valuable insights into the selection of optimal vaccine candidates against *P. falciparum*. Based on our results, we recommend prioritizing conserved merozoite antigens and transmission-blocking antigens. Combining these antigens in multi-stage approaches may be particularly promising for malaria vaccine development initiatives.

**Funding:** Purdue Department of Biological Sciences; Puskas Memorial Fellowship; National Institute of Allergy and Infectious Diseases (U19AI089680).

**Research in context:** *Evidence before this study:* Decades of research on the most virulent malaria parasite, *Plasmodium falciparum*, have yielded multiple antigen candidates of pre-erythrocytic, blood-stage, and transmission-blocking vaccines in varying stages of development from preclinical development to more advanced clinical trials. The malaria vaccine, RTS,S/AS01, which was constructed using the C-terminal and NANP repeat region of the Circumsporozoite Protein (*CSP*) from the African reference strain 3D7, was approved and recommended for use in 2021. However, the vaccine’s lower efficacy is likely a result of the genetic polymorphism of the target antigen shown by studies on natural variation in *CSP*. Similarly, another more recent pre-erythrocytic vaccine, R21/Matrix-M, showed great promise in clinical trials and was recommended in late 2023 by the WHO for use for prevention of malaria in children, but is also multi-dose and *CSP*-based. To maximize vaccine efficacy, it would be more strategic to first understand diversity and variation of antigens across the three types of vaccine classes, targeting various stages of the *P. falciparum* life cycle. Previous studies have reported analyses of vaccine candidate antigens but were mostly limited to pre-erythrocytic and blood-stage antigens, with less focus on transmission-blocking antigens. These studies revealed that most of the pre-erythrocytic and blood-stage antigens are of high diversity due to balancing selection, posing challenges for vaccine design to encompass the antigenic variation. A search conducted on PubMed on April 1, 2024, for relevant published research which used the terms “malaria vaccine”, “*Plasmodium falciparum*” [not “*vivax*”], “selection” and “diversity” yielded 48 studies between 1996 and the present day, with only 14 published studies in the past 3 years. This emphasizes the need for more studies assessing genetic diversity and selection of potential *P. falciparum* vaccine candidates to aid in more effective vaccine development efforts. A similar search with the terms “transmission-blocking vaccine”, “malaria”, “*Plasmodium falciparum*”, not “*vivax*”, “selection” and “diversity” without any date or language restrictions revealed three relevant studies. This warrants future studies to explore transmission-blocking vaccines in this context.

*Added value of this study:* By comparing the genetic and structural analyses of transmission-blocking antigens with pre-erythrocytic and blood-stage antigens, we identify promising *P. falciparum* vaccine antigens characterized by their conservation with low balancing selection and the presence of infection/transmission-blocking epitopes, which are essential for informing the development of new malaria vaccines. This comprehensive workflow can be adopted for studying the genetic and structural variation of other *P. falciparum* vaccine targets before developing the next generation of malaria vaccines for effectiveness against natural parasite populations.

*Implications of this study:* Our suggested strategies for designing malaria vaccines include two possible approaches. We emphasize the development of a multi-stage vaccine that combines critical components such as anti-merozoite (*Rh5*) and transmission-blocking antigens (*Pfs25*, *Pfs28*, *Pfs48/45*, *Pfs230*). Alternatively, we suggest the creation of transmission-blocking vaccines specifically targeting *Pfs25*, *Pfs28* and *Pfs48/45*. These innovative approaches show great potential in advancing the development of more potent and effective malaria vaccines for the future.

## Introduction

For preventing and controlling many infectious diseases, vaccines are vital tools that increase immunity, leading to reduced disease severity, incidence, and transmission. For malaria, a vector-borne disease caused by *Plasmodium* species parasites, vaccine strategies can be characterized by the three main parasite life-cycle stages that they target (Fig. 1). Firstly, pre-erythrocytic vaccines target sporozoites inoculated by *Anopheles* mosquitoes into the human skin before they reach the liver or infected hepatocytes, with the aim of preventing initial infection^1,2^. Some pre-erythrocytic subunit vaccine candidates^3,4^ for *P. falciparum* include the first licensed malaria vaccine RTS,S/AS01^5^, low-dose R21/Matrix-M vaccine (currently approved for use in some countries^6^), and the full-length *CSP* ^7^. Blood-stage vaccines predominantly target merozoites or infected erythrocytes to prevent replication and clinical illness, and some candidates include *AMA1*, *MSP1*, and *Rh5* ^3,8^. Transmission-blocking vaccines target antigens expressed in the gametes and ookinete stages in the mosquito midgut; this vaccine type induces antibodies in humans that impair the parasite’s development inside the mosquito and thus prevent subsequent transmission. Some examples of transmission-blocking vaccine candidates are *Pfs25*, *Pfs230*, and *Pfs48/45* ^3,9,10^.

**Figure 1.**
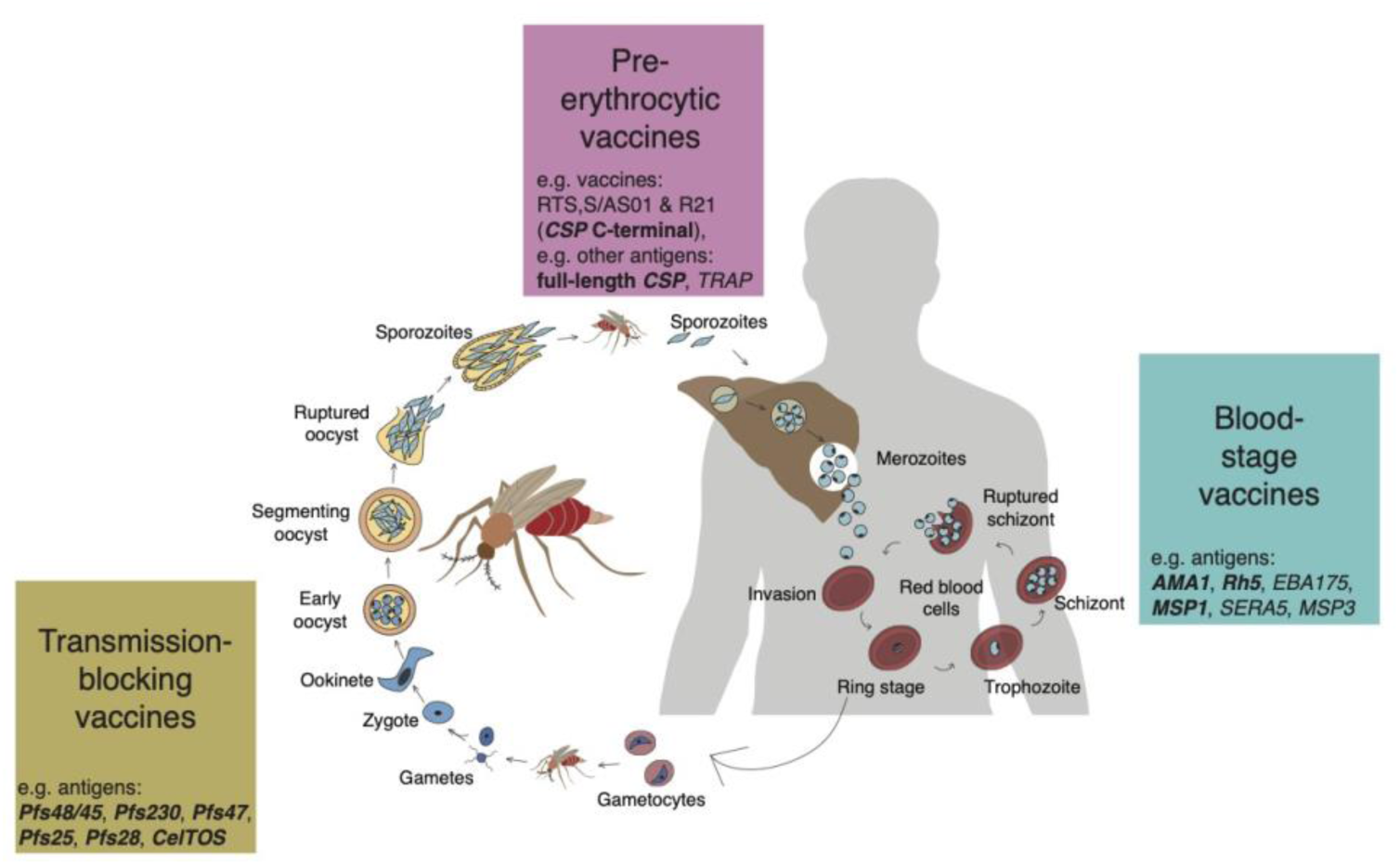
*Plasmodium falciparum* life cycle in the *Anopheles* mosquito vector and human host and corresponding targets for malaria vaccine development. Vaccine strategies are broadly grouped by the *P. falciparum* life stage they target and include the categories: pre-erythrocytic, blood stage, and transmission-blocking. The pre-erythrocytic class of vaccines usually include candidates that target sporozoites or infected hepatocytes to prevent clinical initial infection and stop malaria parasites from re-entering the bloodstream. The blood stage vaccines target the parasite during replication in red blood cells in the human. Lastly, transmission-blocking vaccines target sexual and/or sporogonic development stages and aim to induce antibodies in humans that impair the parasite’s development inside the mosquito midgut and thus subsequently prevent the transmission of sporozoites to humans. The antigens shown in bold were included in this study.

To date, among the numerous preclinical and clinical trials of these classes of vaccines, RTS,S and R21 are the only two malaria vaccine licensed and recommended by the WHO. The former is approved for widespread use among children in sub-Saharan Africa and other regions with moderate to high *P. falciparum* malaria transmission^5,11–14^ and the latter by regulators in Ghana, Nigeria, and Burkina Faso^6,15^. While this is a tremendous breakthrough for the field considering the complexity of the parasite life cycle and the correspondingly large number of protein-coding genes (>5,000) in the *Plasmodium* genome^16^, there is a wealth of vaccine targets^17–20^. Despite the large number, about 60% of the genes code for putative proteins^21^ and have unknown function^16,22^, which poses an enormous challenge, and many of these 5300+ protein coding genes do not make attractive vaccine targets due to expression patterns, localization, high genetic variability, etc., and candidates must be experimentally validated for vaccine development^23^. Yet, despite decades of effort, noticeably little progress past early phase clinical trials for many of these other targets^24,25^. Furthermore, the RTS,S vaccine shows only modest efficacy ranging from 30% to 40% that wanes with time and requires four doses to reduce severe malaria by 30%^12,26–28^. This low efficacy has been attributed to the high genetic diversity and many polymorphic residues of the target antigen, *CSP* ^12,24,29^. Notably, RTS,S was developed using a standard laboratory strain (*P. falciparum* 3D7), which originates from Africa but is not highly representative^30^ and has low frequency within many parasite populations^29,31–33^. This underlines the need to consider *CSP*’s functional importance as well as the natural variation in the population, especially in epitope regions^25,34^, when designing a vaccine. In recent years, vaccines based on regions of conserved or semi-conserved epitopes are emerging in other important yet antigenically diverse pathogens such as influenza virus and meningococcus^35–37^.

With the release of thousands of publicly available *P. falciparum* whole genome sequences (WGS), it is possible to study the natural genetic and functional variation of targets for malaria vaccine candidate antigens, yet, to date few studies have done so^34,38–40^. Two recent studies reported comprehensive analyses of vaccine candidate antigens^25,41^, but were focused primarily on pre-erythrocytic and blood-stage antigens, with limited inclusion of transmission-blocking antigens. Here, we aimed to characterize the natural genetic and structural variation of ten *P. falciparum* vaccine antigens spanning the three different vaccine classes, including transmission-blocking antigens. We focused on Africa which carries the heaviest burden of malaria, and augmented parasite genomic data from Zambia, where malaria continues to be endemic and a major public health with over 8 million cases and 1300 deaths reported in 2022^42^.

## Methods

### Ethics Statement

The parents or legal guardians provided parental permission for study participants and this study was conducted with the approval of the Biomedical Research Ethics Committee from the University of Zambia (Ref 011-02-18) and from the Zambian National Health Research Authority.

### Sample Selection and Processing

This work is a secondary analysis of a larger parent study focused on understanding malaria transmission across Zambia and establishing a baseline for parasite genetic metrics for this country that are informative about transmission intensity, genetic relatedness between parasites, and selection^43^. Briefly, dried blood spot (DBS) samples were collected as part of the 2018 Zambia Malaria Indicator Survey (MIS), which used a nationally representative two-stage stratified clustering sampling strategy to obtain samples across Zambia for the estimation of malaria prevalence^44^. The 2018 MIS included DBS samples from children under the age of 5 from 179 standard enumeration areas from across ten provinces, with high transmission provinces oversampled^44^. Due to undersampling in the low transmission provinces (Central, Lusaka and Southern), we augmented the DBS samples from the Zambia 2018 MIS with DBS samples from Southern Province collected in the community surveillance program from the Southern and Central Africa International Centers of Excellence for Malaria Research (ICEMR)^45^. These additional samples temporally matched the MIS (from 2018-2019) and were collected as part of a longitudinal cohort study conducted to understand sources of focal transmission from an area with an expected relatively high number of incident malaria cases.

In total we included 459 PET-PCR positive *P. falciparum* samples from seven provinces^43^ and additional 28 PCR positive *P. falciparum* samples from Southern Province for sequencing. The samples were processed for genomic DNA extraction as previously described^43,46^ and then genomic library preparation, parasite enrichment and sequencing were carried out.

### Whole-Genome Sequencing

To selectively enrich and sequence whole *P. falciparum* genomes from DBS we used a 4-plexed hybrid capture method previously described^43^. Genomic library preparation, hybridization capture, and sequencing was conducted at the Yale Center for Genomic Analysis (YCGA). Samples were sequenced using 101 bp paired-end read sequencing on an Illumina NovaSeq 6000 with a target of 30 million reads per sample.

### Acquisition of Additional Genomic Data

To contextualize Zambian *P. falciparum* genome diversity within Africa, we included and analyzed 781 publicly available *P. falciparum* WGS data from 5 countries across West, Central, and East Africa (Democratic Republic of the Congo, Ghana, Guinea, Malawi, and Tanzania) from the MalariaGEN Pf3k project release 5.1 (https://www.malariagen.net/data/pf3k-5) and the MalariaGEN *P. falciparum* Community Project (http://www.malariagen.net/resource/16). Raw Fastq files were downloaded from SRA using pysradb (https://github.com/saketkc/pysradb)^47^ and processed in the same way as the newly sequenced WGS.

### Read Mapping and Variant Identification

We identified *P. falciparum* genomic variation from whole genome sequence data using a bioinformatics pipeline previously described^48^. Briefly, the Illumina paired-end reads were trimmed for low-quality bases (Phred-scaled base quality <30) and mapped to the *P. falciparum* 3D7 reference genome (ftp://ftp.sanger.ac.uk/pub/project/pathogens/gff3/2015-08/Pfalciparum.genome.fasta.gz) using BWA-MEM v0.7.17^49^. Aligned reads for the 1248 samples (487 from Zambia and 781 from Pf3k) were marked for duplicates and sorted using Picard Tools 2.20.8. For variant calling only samples with >50% of the regions spanning the selected antigens (Table 1) with >5X read coverage were included, resulting in a total of 1092 *P. falciparum* WGS samples. Variants were called using GATK v4.1.4.1 following best practices (https://software.broadinstitute.org/gatk/best-practices)^50,51^. We applied GATK Base Quality Score Recalibration using default parameters and using variants from the *P. falciparum* crosses 1.0 release as a set of known sites (ftp://www.malariagen.net/data/pf-crosses-1.0)^52^. GATK HaplotypeCaller in GVCF mode was used to call single-sample variants (*ploidy* 2 and *standard-min-confidence-threshold for calling* = 30), followed by GenotypeGVCFs to genotype the cohort. We restricted our analyses to variants from the 14 nuclear chromosomes only, excluding variants from telomeric and hypervariable regions (ftp://ngs.sanger.ac.uk/production/malaria/pf-crosses/1.0/regions-20130225.onebased.txt). We further removed variants with VQSLOD <0 and SNPs with missingness >0.2. After variant filtering, we scored 426,757 genome-wide SNPs across the 1092 samples. Variants were functionally annotated with SnpEff (version 4.3t) for genomic variant annotations and functional effect prediction^53^.

**Table 1.**
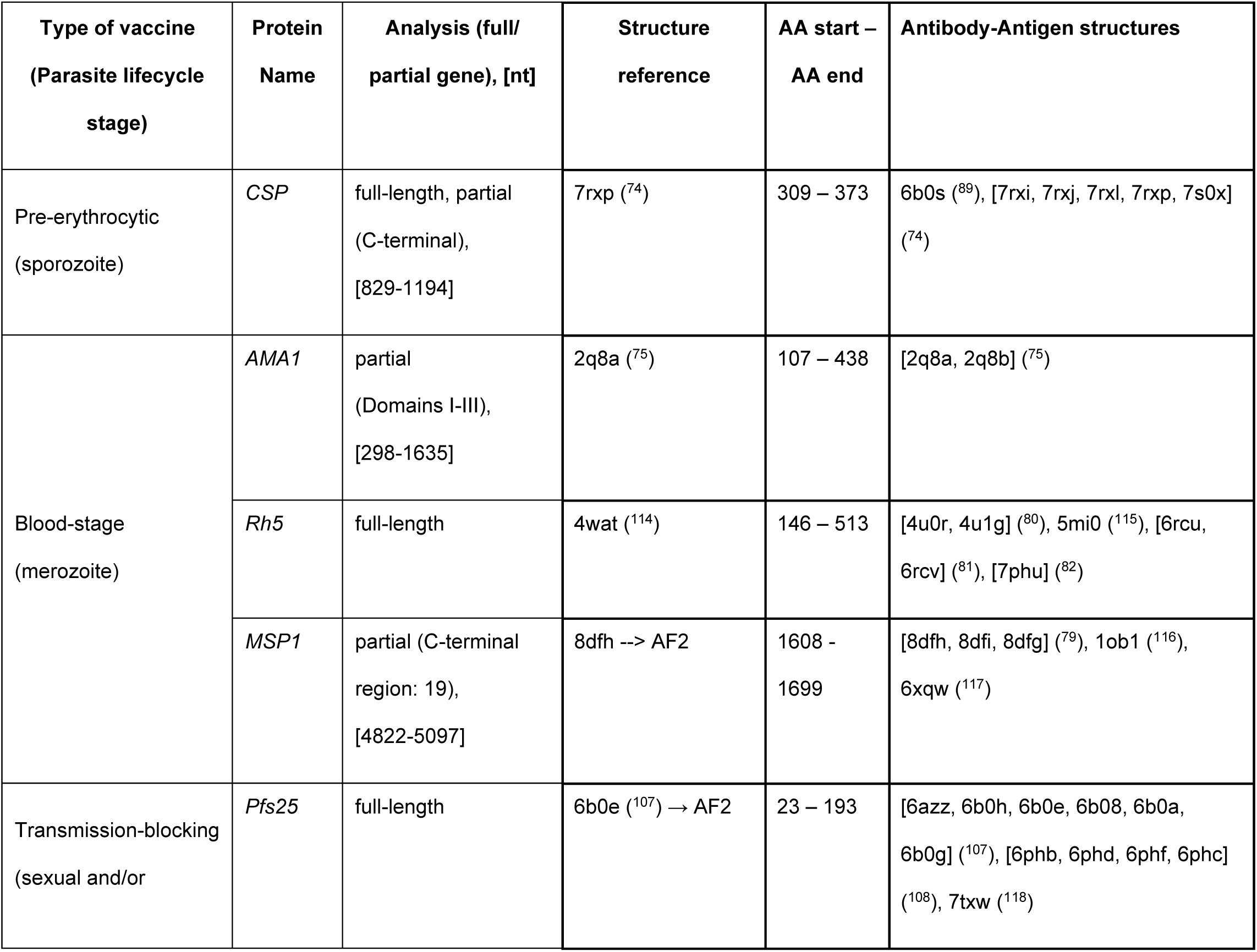
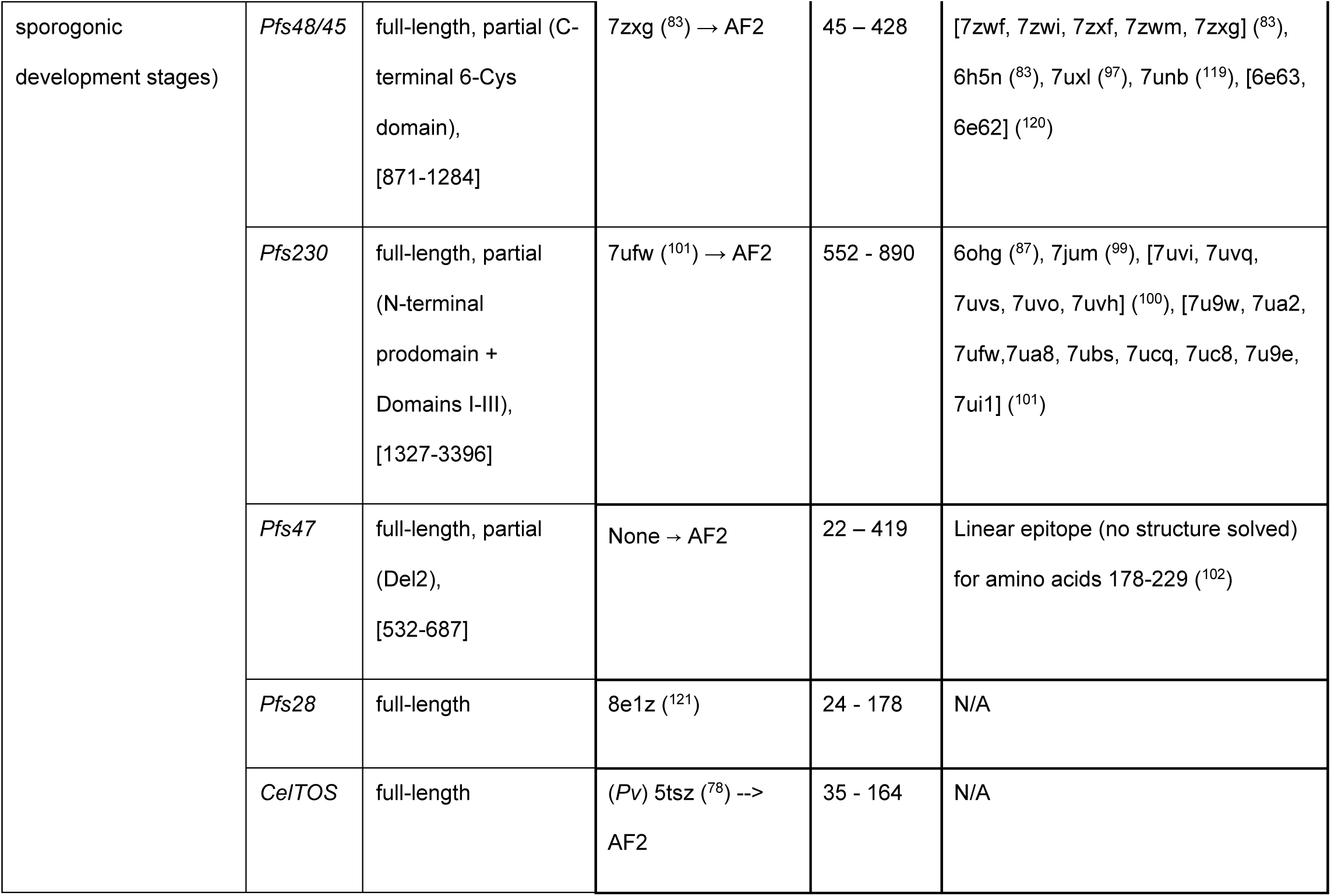
Vaccine candidate antigens. *P. falciparum* antigens included in this study, with respective type of vaccine, lifecycle stage, full or partial gene used in analyses from NCBI gene database, and 3-D structure reference and antibody-antigen structures used in analyses from Research Collaboratory for Structural Bioinformatics (RCSB) database. The gene information of the proteins is provided in Table S1. All are relevant key candidates from malaria vaccine trials and are functionally important for malaria biology as they target different stages of the *Plasmodium falciparum* life cycle (See Figure 1). AF2 represents AlphaFold solved structures. “→” indicates we used AlphaFold to resolve loop regions instead of the solved structure (see Methods for details).

### Multiplicity of Infection

To ensure high quality haplotype reconstruction, we first determined the multiplicity of infection (MOI) for each sample using within-sample F statistic (*F_WS_*)^54^ implemented in the R moimix package (available at https://github.com/bahlolab/moimix). *F_WS_* scores infections on a continuous scale of multiplicity, with infections having an *F_WS_* > 0.95 considered clonal (MOI=1), those with *F_WS_* < 0.95 and > 0.8 containing two strains (MOI=2), and those with *F_WS_* < 0.8 harboring more than two strains (MOI>2). *F_WS_* metric derived from the 426,757 genome-wide SNPs were used to stratify samples into single infections, two infections or more than two infections.

### Selection of Vaccine Candidate Antigens

Antigens were selected based on their inclusion^14^ in either two licensed and recommended malaria vaccines (e.g. *CSP*)^28^, clinical development in Phase I (e.g. *AMA1*, *Rh5*, *MSP1*, *Pfs25*, *Pfs48/45*, and *Pfs230*)^3^, clinical development in Phase II (e.g. *Rh5* and *Pfs230*)^55–57^, or their recognition as a promising vaccine target (e.g. *Pfs47*, *Pfs28*, and *CelTOS*)^58,59^. The ten selected antigens encompassed all parasite life cycle stages (Table 1 and Fig. 1), and genomic information and coordinates are provided in Table S1.

### Haplotype Reconstruction

Vaccine target regions (Table S1) were extracted from the filtered VCF file (VQSLOD <0) in BED format using vcftools (version 0.1.16). Haplotypes were reconstructed using a custom code available at https://github.com/giocarpi/Pf_capture_div_code, which converted the VCF file to FASTA format, incorporating all variants (SNPs and INDELs) within coding sequences (CDS) relative to the *P. falciparum* 3D7 reference gene sequences. For a given variant position in a sample, a minimum read depth was required to be 5 for samples of MOI=1 and a minimum read depth of 10 for samples of MOI=2. A minimum minor allele proportion for a heterozygous call to be made was set to 0.2. If the mean frequency of minor alleles was below 0.4, these minor alleles constituted a minor haplotype, which would be included in the following analyses; otherwise, the minor haplotype was not retained (according to paper^25^ and code). Samples failing read depth criteria were excluded from downstream analysis, as were samples containing one or more ‘N’ nucleotides.

### Genetic Diversity Analysis

The software DNA Sequence Polymorphism (DnaSP) version 6.12.03 was used to calculate the number of unique haplotypes (h) of each respective vaccine candidate antigen in each country, the number of polymorphic sites (S), and the average number of pairwise nucleotide differences (k) between samples from within and between countries^60^. Furthermore, we calculated the haplotype diversity (Hd) for each antigen, with 1 representing a population in which every haplotype is unique and 0 equating a population where each haplotype is identical. Nei’s nucleotide diversity statistic^61^ (π) within each country and among all samples was also calculated, which represents the average number of nucleotide differences between haplotypes drawn from the same population. *Tajima’s D* statistic values^61–63^ were calculated using Molecular Evolutionary Genetics Analysis (MEGA X) version 10.1.7 to measure selection (*Tajima’s D* values that are approximately zero occur in neutral conditions, >1 indicates balancing selection, and <1 represents purifying selection). Gaps in the multiple sequence alignments were treated as complete deletion. To assess the genealogical relationships between the haplotypes found in Pf3k from African countries and those from Zambia for each vaccine candidate antigen of interest, we constructed haplotype networks based on the method by Templeton, Crandall, and Sing (TCS) using PopArt version 1.7^64,65^.

### Structural Analyses

Protein structures used for the downstream analyses were either selected from existing solved ones or generated from AlphaFold2^66^. PDB structures were retrieved from the Research Collaboratory for Structural Bioinformatics (RCSB) database on 5/23/2023. The final solved structures to be used for each protein were selected based on three criteria: 1) the structure was based on *P. falciparum* 3D7 reference gene sequences; 2) most complete in terms of gene coverage; 3) resolved as many residues with polymorphisms as possible. Often, the SNP-containing residues occurred in solvent-exposed loop regions that were not resolved in published Protein Data Bank (PDB) structures. In these cases (i.e., *AMA1*, *Rh5*, *Pfs230* (D1, D2)*, Pfs25, MSP1_19_*) and the case of no solved structures for *P. falciparum* (i.e., *Pfs47, CelTOS*), we assessed the qualities of AlphaFold2’s predictions of these structures, that is whether the RMSD is small or < 3 Å^67,68^ (Fig. S3). *Pfs230* (D1, D2), *Pfs25, Pfs4845,* and *MSP1_19_* were successfully folded by AlphaFold2, agreeing with the native fold at or below 2 Å Root Mean Square Deviation (RMSD). *Rh5* prediction was unable to meet this threshold, therefore we proceeded with the most complete solved structure of *Rh5* available. *AMA1* structures were missing domain III, as well as missing resolution of many residues in a loop of domain II (residues 351–389). AlphaFold2 was able to confidently predict domain III and the overall structure (RMSD 0.36), but the missing loop residues of domain II were of low confidence, and the placement of the loop disagreed with the resolved residues of this loop in solved structures. For this reason, we proceeded with an AlphaFold2 model of *AMA1* including domain III but excluding the low confidence loop region of domain II.

*Pfs47* had no deposited structures. Hence, it was predicted entirely with AlphaFold2 with good confidence (Fig. S3). *CelTOS* had no deposited structures for *P. falciparum,* but the *P. vivax* homolog has been experimentally solved. We assessed a structure modeled with AlphaFold2 for *P. falciparum CelTOS* and verified its fit to *P. vivax* (RMSD 1.59), trimming off residues outside the solved *P. vivax* structure.

AlphaFold2 structures were predicted on 3D7 reference sequences, utilizing the ColabFold^69^ pipeline (https://colab.research.google.com/github/sokrypton/ColabFold). Multiple sequence alignments were performed with MMseqs2 searching through UniRef+Environmental datasets. For each protein, three different structure models were run with six recycles and early stops at 100 PLDDT. Templates and AMBER relaxation were set to “off”. Models of rank one were chosen as final structures. For predicted structures with solved references that needed loop or domain modeling, an agreement between predicted and solved reference of 2 Å RMSD or less was deemed suitable for downstream analysis. For predicted structures with no solved reference, a convergence of three models to a structure within 2 Å RMSD between three models was assessed as a confident prediction model.

All the available solved antibody-antigen structures of seven of ten vaccine target proteins were obtained from RCSB (as of 7/12/23). Using a custom R script, we extracted interface residues between the antibody and target protein that occur within 5 Å of each other. *Pfs47* lacks antibody-antigen structures, so we have included a known linear epitope (see Table 1). These interface residues were sorted into bins for visualization purposes at 0, 1, 2, or 3+ antibody interfaces, to represent common epitopes vs. infrequent and non-targeted residues. We consolidated identical antibodies with multiple structures into a single record. This ensures that the count of epitopes involving each interface residue only includes unique antibody contacts.

### Calculation of 3D population genetics measures

To calculate spatially derived nucleotide diversity (π*) and Tajima’s D (D*) values, we utilized the BiostructMap Python^70,71^ package, installed from https://github.com/andrewguy/biostructmap. The package provides a method for mapping DNA sequence data to residues on protein structures, defining a spatial neighborhood of the residue, and calculating the statistics using a three-dimensional (3D) sliding window approach. In our study, we selected a 3D window with a radius of 15 Å, which represents the maximum surface of an antibody-antigen binding site^72^. We chose not to apply a relative solvent accessibility (RSA) filter and included all the residues within the radius of 15 Å, because polymorphisms at buried residues could have structural effects on an epitope, and thus should be reflected in calculated diversity and selection pressure.

## Results

### Enhancing *P. falciparum* genomic diversity from African countries

We augmented 241 *P. falciparum* whole genome sequences previously reported^43^ with an additional 84 genomes from Zambia, a malaria endemic country in South-Central Africa currently lacking *P. falciparum* genomic representation in public databases. Combining these data with publicly available WGS sequences from the MalariaGEN *P. falciparum Community Project Consortium*^73^ (http://www.malariagen.net/resource/16), WGS data of 1248 *P. falciparum* samples from 6 African countries were processed to obtain gene sequence haplotypes for ten antigens, with the final data set consisting of 1092 samples after quality filtering (Table 1, Table 2) for population genetic analyses. To ensure high-quality haplotype reconstruction, the multiplicity of infections (MOI) was determined using *F_ws_*^54^, and samples with a high multiplicity of infections (MOI>2) were removed. Overall, 44.2% (483) of 1092 African field samples were found to harbor more than two infections, with Malawi having the highest percentage (50.8%), followed by Zambia (48.9%) (Fig. S2). Moreover, 17.6% of samples (192) had MOI=2 and 38.2% (417) were characterized as MOI=1 (Fig. S2). Additionally, there was a mean of 457 (range:196-579) sequences per antigen when considering full-length genes for analysis, and a mean of 591 (range: 555-623) sequences per antigen when considering partial genes for those with specific regions of interest related to vaccine targets (Table 2).

**Table 2.**
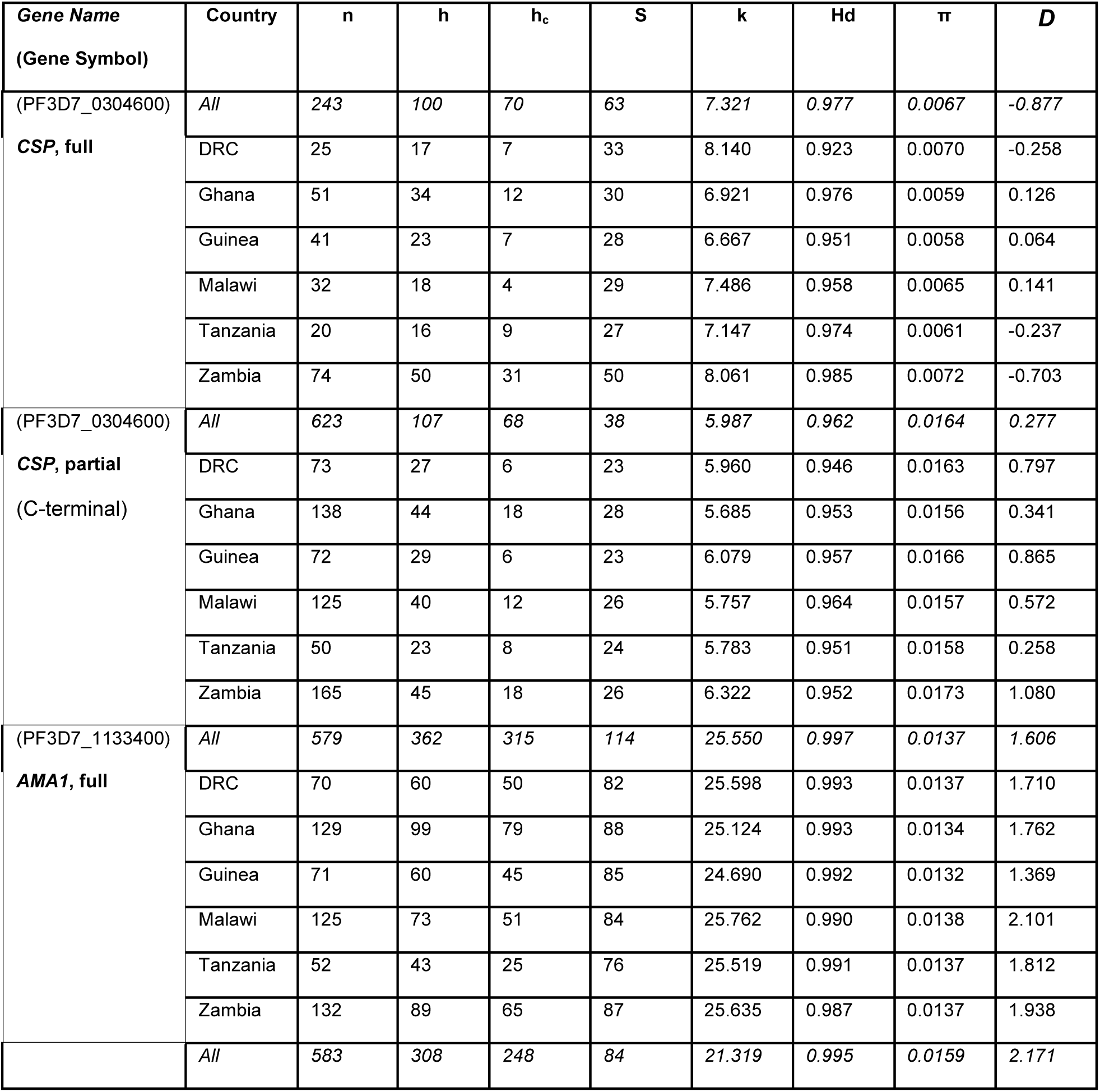
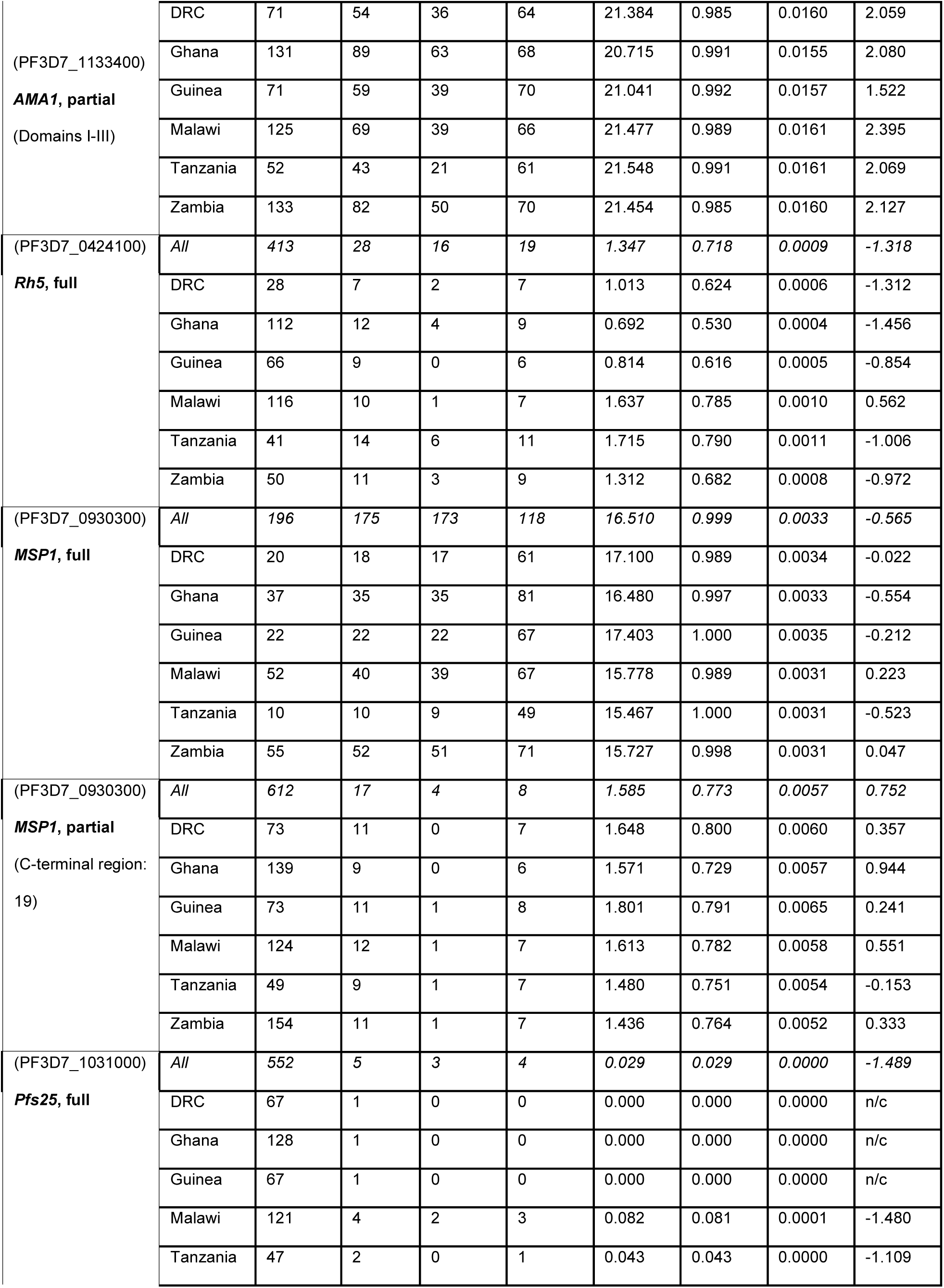
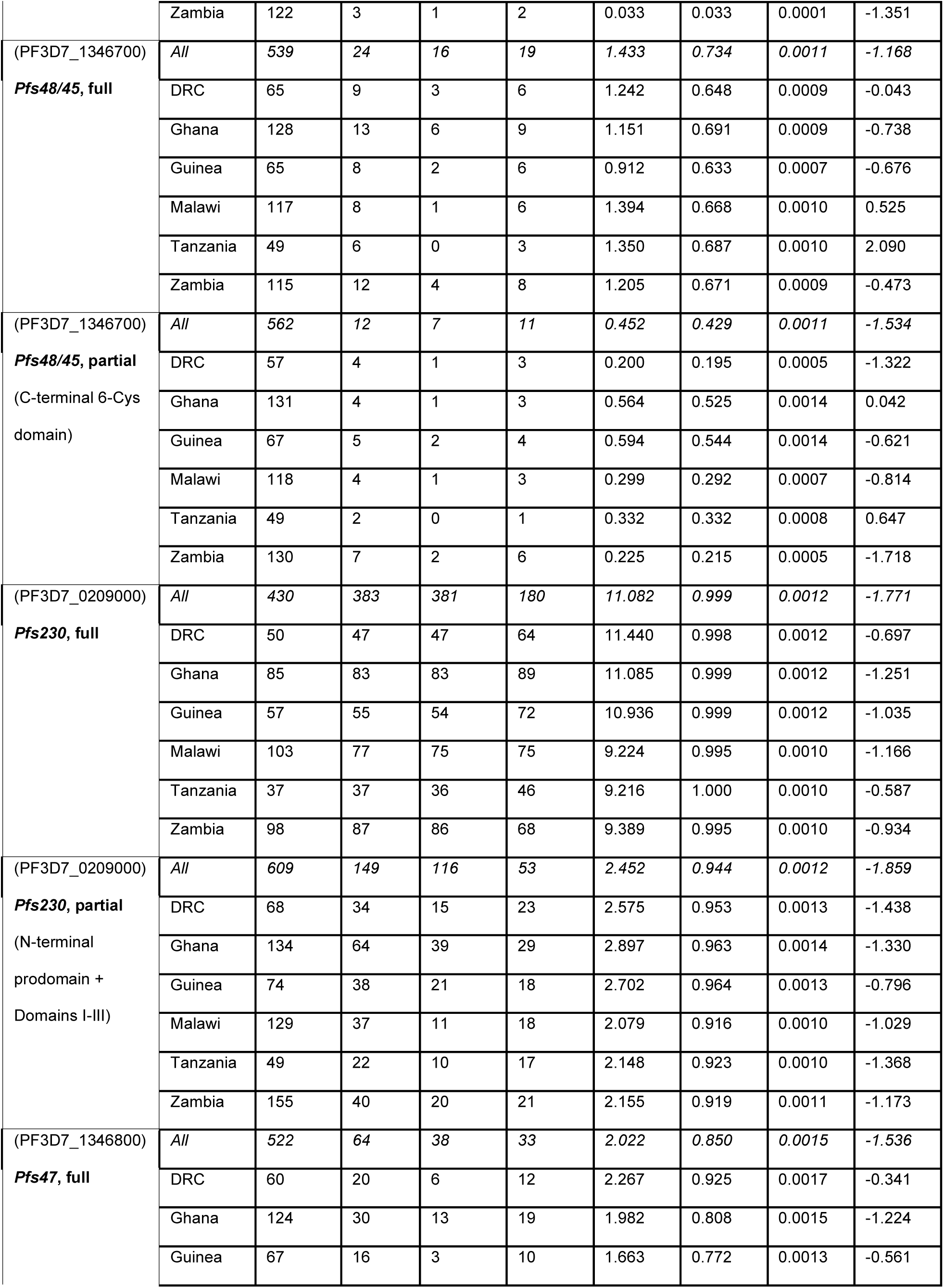
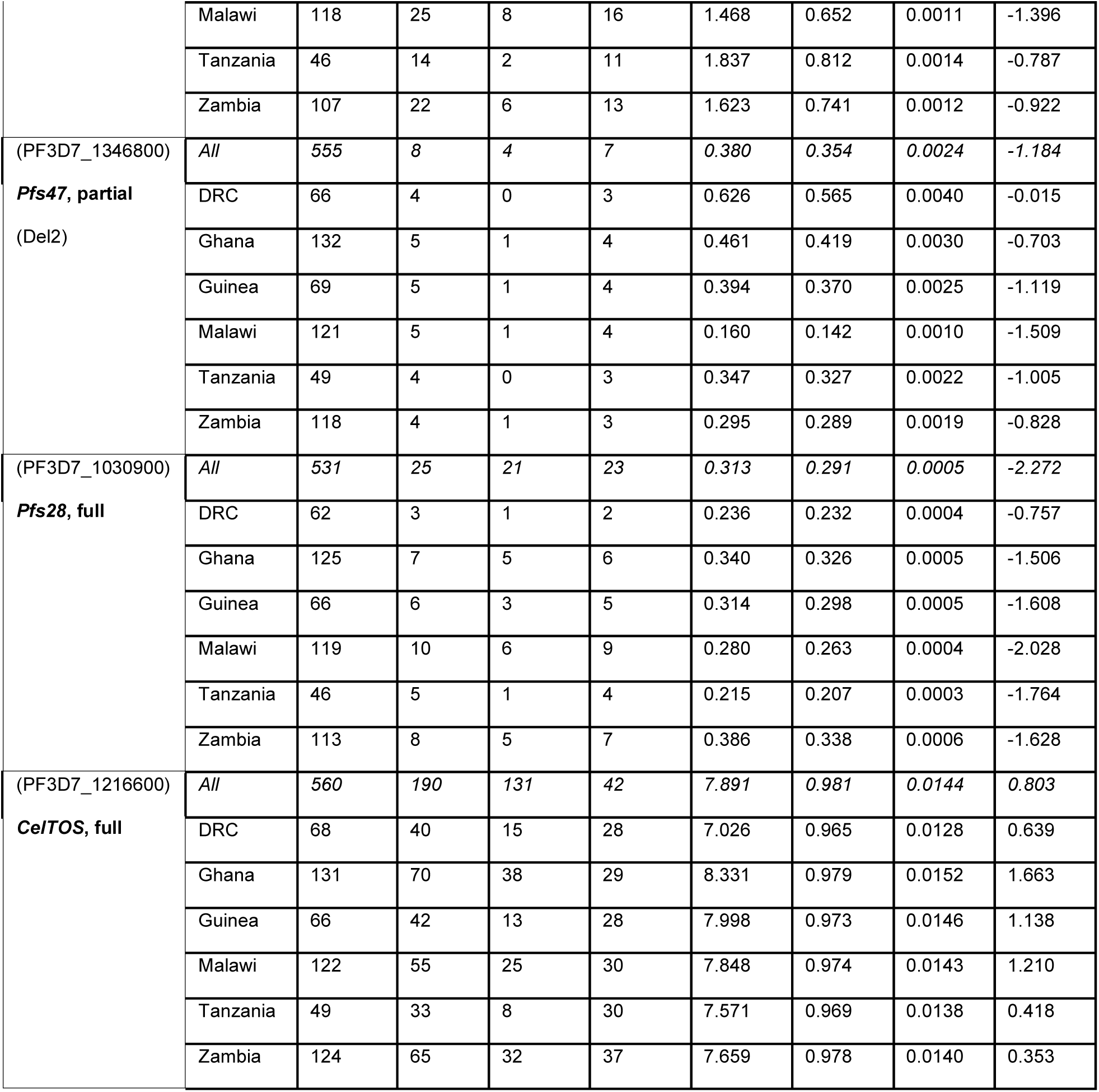
Diversity Statistics Results. Summary of the samples from Africa included in our analyses per gene. n=number of sequences, h=number of unique haplotypes, h_c_=number of unique haplotypes from a respective country alone, S=number of polymorphic (segregating) sites, k=average number of pairwise nucleotide differences, Hd=haplotype (gene) diversity, π=nucleotide diversity, *D*=Tajima’s neutrality test statistic

### Antigen haplotypes lack geographic structure

To assess the genealogical relationships amongst sequence haplotypes obtained from our newly derived Zambian sequences and those from other African countries, we used TCS network analysis for the ten antigens. Full length genes, as well as known vaccine target regions, were examined and compared (Fig. 2 and 3). In general, haplotypes did not exhibit clustering, irrespective of vaccine antigen type and geographic origin. This observation held true whether the analysis considered the entire gene or specific functionally relevant domains, suggesting a lack of geographic structure. (Fig. 2 and 3).

**Figure 2.**
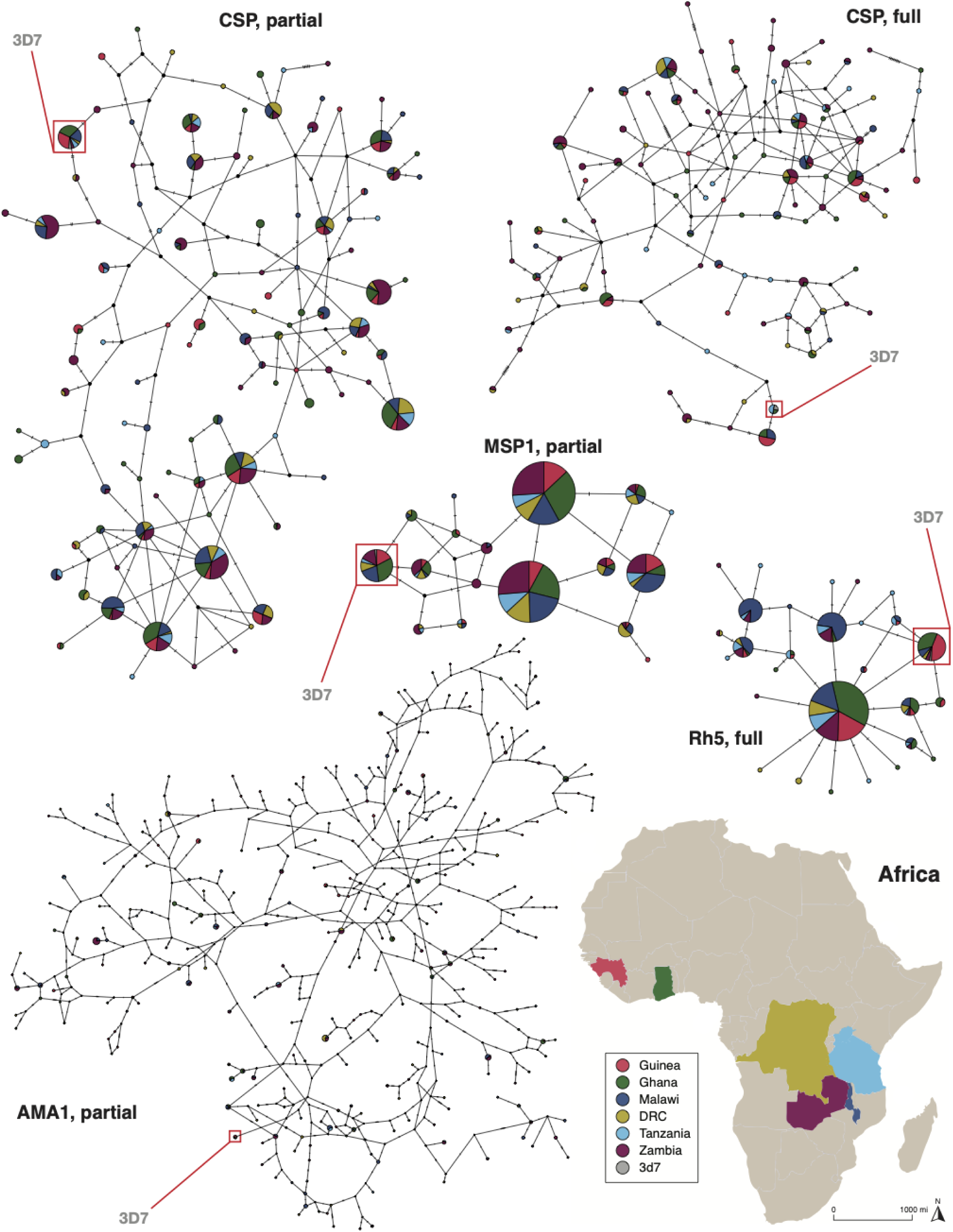
Haplotype network for malaria vaccine antigens (pre-erythrocytic and blood-stage proteins). Templeton, Crandall, and Sing (TCS) networks illustrate the diversity of the selected antigens. Circles represent unique nucleotide haplotypes and are scaled according to the frequency with which the haplotype was observed. The number of mutation differences that exist between each haplotype is depicted by the number of hatch marks on the network branches. Haplotype colors match the geographic origin of the samples depicted on the map (see map legend). The vaccine strain 3D7 (grey color, surrounded by a red box) is included for reference in each respective haplotype network.

**Figure 3.**
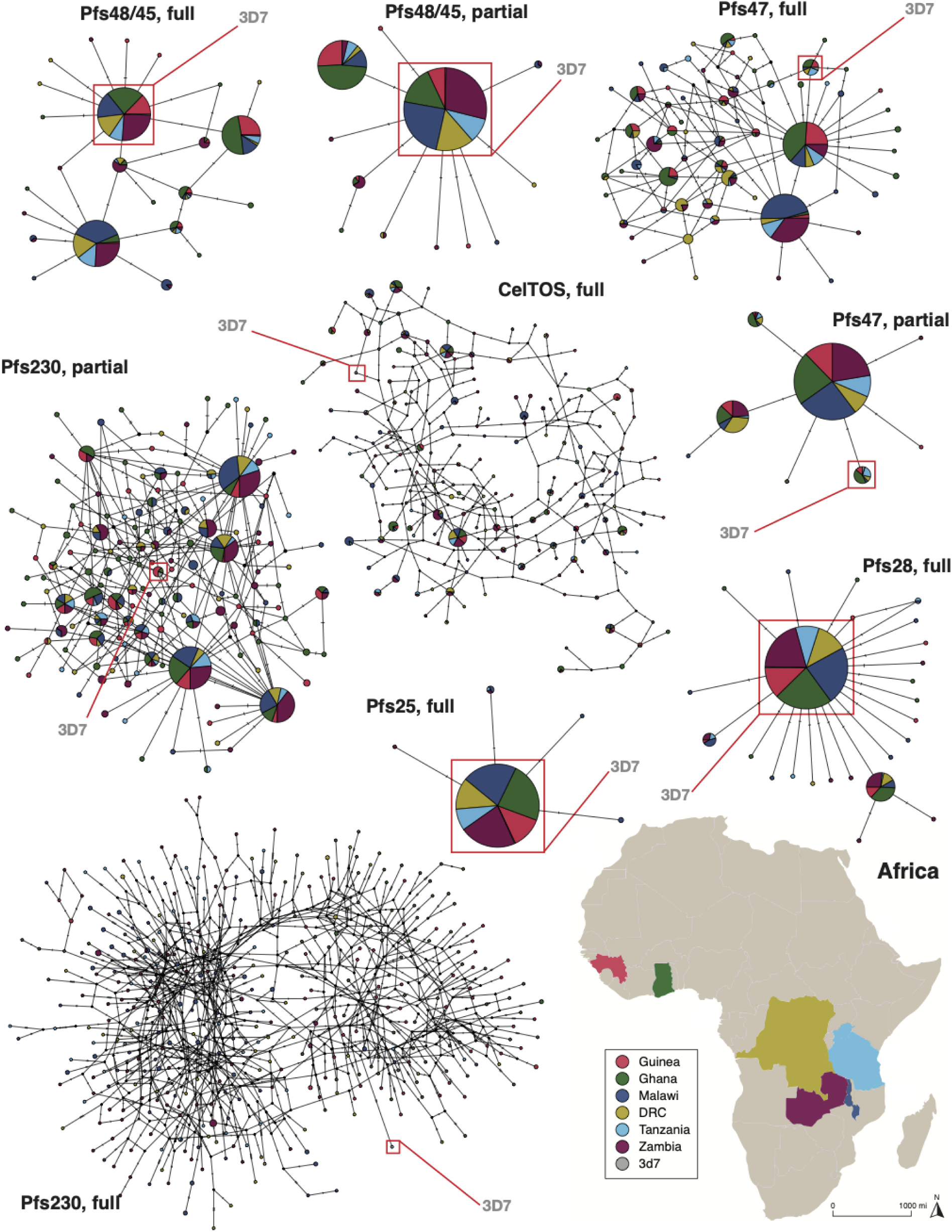
Haplotype network for malaria vaccine antigens (transmission-blocking proteins). Templeton, Crandall, and Sing (TCS) networks illustrate the diversity of the selected antigens. Circles represent unique nucleotide haplotypes and are scaled according to the frequency with which the haplotype was observed. The number of mutation differences that exist between each haplotype is depicted by the number of hatch marks on the network branches. Haplotype colors match the geographic origin of the samples depicted on the map (see map legend). The vaccine strain 3D7 (grey color, surrounded by a red box) is included for reference in each respective haplotype network.

We identified the following percent of unique haplotypes across all countries for individual full-length genes: 41.2% (100/243) for *CSP*, 62.5% (362/579) for *AMA1*, 6.8% (28/413) for *Rh5*, 89.3% (175/196) for *MSP1*, 0.9% (5/552) for *Pfs25*, 4.5% (24/539) for *Pfs48/45*, 89.1% (383/430) for *Pfs230*, 12.3% (64/522) for *Pfs47*, 4.7% (25/531) for *Pfs28*, and 33.9% (190/560) for *CelTOS*. For certain genes, we examined specific regions of interest related to vaccine targets and identified the following percent of unique haplotypes across all countries: 17.2% (107/623) for *CSP* C-terminal, 52.8% (308/583) for *AMA1* (Domains I-III), 2.8% (17/612) for *MSP1* (C-terminal region: 19), 2.2% (12/562) for *Pfs48/45* (C-terminal 6-Cys domain), 24.5% (149/609) for *Pfs230* (N-terminal prodomain and Domains I-III), and 1.4% (8/555) for *Pfs47* (Del2). We further separated unique haplotypes by country for each gene (see Table 2 for full diversity statistics results breakdown). To contextualize this specifically for Zambia, when considering unique haplotypes solely from this country, we observed a modest number across all genes: 44.3% (31/70) for *CSP* full-length and 26.5% (18/68) for *CSP* partial, 20.6% (65/315) for *AMA1* full-length and 20.2% (50/248) for *AMA1* partial, 18.8% (3/16) for *Rh5* full-length, 29.5% (51/173) for *MSP1* full-length and 25% (1/4) for *MSP1* partial, 33.3% (1/3) for *Pfs25* full-length, 25% (4/16) for *Pfs48/45 full-length* and 28.6% (2/7) for *Pfs48/45* partial, 22.6% (86/381) for *Pfs230* full-length and 17.2% (20/116) for *Pfs230* partial, 15.8% (6/38) for *Pfs47* full-length and 25% (1/4) for *Pfs47* partial, 23.8% (5/21) for *Pfs28* full-length, and 24.4% (32/131) for *CelTOS* full-length. The 3D7 vaccine strain haplotype was dominant for antigens *Pfs48/45* (full-length and partial), and *Pfs25* and *Pfs28* (both full-length) (Fig. 3). In contrast, for the more diverse antigens i.e., *Pfs230*, *AMA1*, *CSP* and *MSP1,* the 3D7 vaccine strain haplotype was found at very low frequencies (Fig. 2 and 3).

### Differing signals of selection, haplotype, and nucleotide diversity across antigen types for vaccine development considerations

To understand haplotype diversity and investigate evidence of selection, we derived Nei’s nucleotide diversity (*π*), haplotype diversity (Hd) and *Tajima’s* D statistics for each antigen. When taking full-length genes into consideration, overall, five antigens, *Pfs230*, *AMA1*, *CSP*, *MSP1*, and *CelTOS*, representing genes across the three categories of vaccine targets, exhibited the highest values of haplotype diversity (Hd >0.9) across all parasite populations analyzed (Fig. 4a, Table 2). Two transmission-blocking antigens, *Pfs47* (0.85) and *Pfs48/45* (0.73) showed a moderate-high level of haplotype diversity, with similar levels as those demonstrated by the blood-stage antigen *Rh5* (0.72) (Fig 4a, Table 2). The transmission-blocking antigen *Pfs28* exhibited low haplotype diversity (0.29), and the lowest haplotype diversity was observed for another transmission-blocking antigen, *Pfs25* (0.03) (Fig. 4a, Table 2). When considering antigens with specific regions of interest for vaccine development, the same trend was observed: *AMA1*, *CSP*, and *Pfs230* showed extremely high levels of haplotype diversity (0.99, 0.96, and 0.94, respectively), followed by *MSP1* with high haplotype diversity (0.77), while *Pfs48/45* had moderate haplotype diversity (0.43), and finally followed by *Pfs47* with the lowest haplotype diversity (0.35) (Fig. 4a, Table 2). Furthermore, across the parasite populations, when examining all of the full-length genes, we detected the highest levels of nucleotide diversity for *CelTOS* (0.0144) and *AMA1* (0.0137) (Fig. 4b, Table 2). The next highest levels of nucleotide diversity were shown for *CSP* (0.0067), *MSP1* (0.0033), and this was followed by similar levels observed for *Pfs47* (0.0015), *Pfs230* (0.0012), and *Pfs48/45* (0.0011) (Fig. 4b, Table 2). These values were tenfold higher than the nucleotide diversity observed for genes *Rh5 and Pfs28* (0.0009 and 0.0005, respectively, and one hundred-fold lower than the nucleotide diversity observed for *Pfs25* of 0.00001 (Fig. 4b, Table 2). When considering partial regions of interest for vaccine development for the included genes, a similar pattern was observed; namely, *CSP* and *AMA1* had the highest nucleotide diversity (0.016 and 0.014, respectively), while *MSP1*, *Pfs47*, and *Pfs230* had tenfold lower nucleotide diversity (0.0057, 0.0024, and 0.0012), and *Pfs48/45* had the lowest recorded nucleotide diversity of 0.0011 among the genes for which partial regions were studied (Fig. 4b, Table 2). When examining specimens from Zambia alone, haplotype and nucleotide diversity aligned with the observed ranges in the parasite populations across the three categories of vaccine targets.

**Figure 4.**
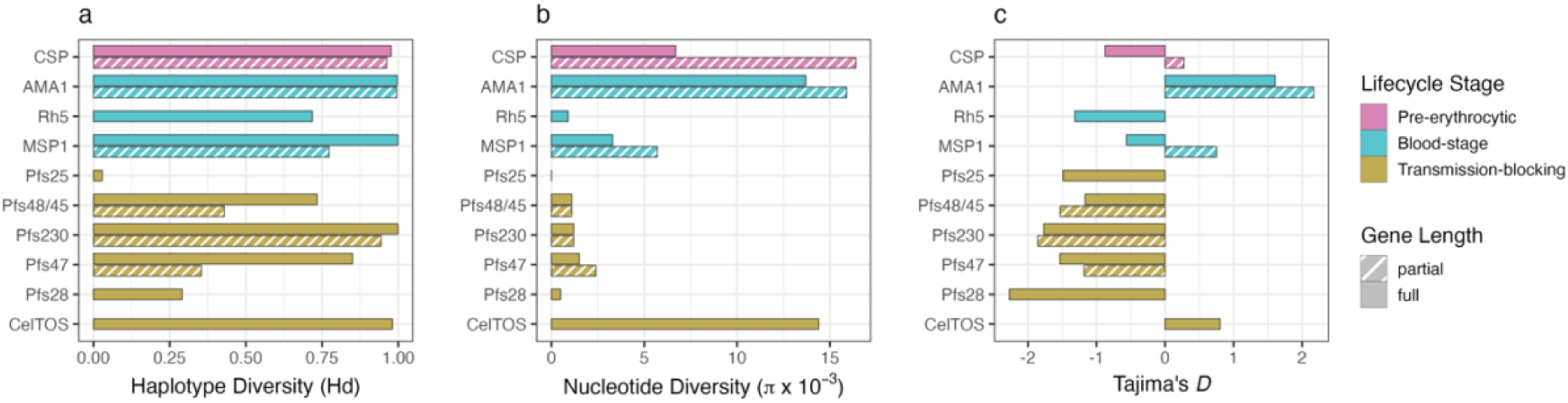
Results of haplotype diversity, nucleotide diversity, and Tajima’s D values for each antigen across all countries included in this study. The horizontal barplot depicts the calculated values for the specified diversity parameters for each antigen across the different parasite populations. Three diversity parameters were included: haplotype diversity (panel a) and nucleotide diversity (panel b) (calculated using DNAsp), and Tajima’s D statistic (panel c) (calculated using MEGA). Colors are selected to show the lifecycle stage of the parasite of the respective antigen (see legend for specific colors) and also correspond to the colors depicted in Fig. 1 of the *P. falciparum* parasite life cycle stages. The fully shaded-in bars indicate full-length genes, while the dashed bars denote partial length genes for genes with specific regions of interest for vaccine development.

To assess SNP neutrality across genes, we conducted *Tajima’s D* statistical test, comparing average pairwise differences between samples and the total number of segregating sites (Fig. 4c, Table 2). *Tajima’s D* values of 0 indicate neutrality in constant-size populations, negative values suggest purifying selection, and positive values suggest balancing selection. When examining full-length antigens, *AMA1* showed the strongest signature of balancing selection, with *CelTOS* displaying neutrality and weaker signs of balancing selection, *CSP* exhibiting neutrality with signs of purifying selection, and *MSP1* similarly showing neutrality with signs of purifying selection (Fig. 4c, Table 2). Conversely, *Pfs28*, *Pfs230*, *Pfs47*, *Pfs25*, *Rh5, Pfs48/45* (in decreasing order) showed clear signatures of purifying selection (Fig. 4c, Table 2). When inspecting antigens with partial regions of interest for vaccine development, we found that *AMA1* still demonstrated strong balancing selection, while *MSP1* and *CSP* showed moderate signs of balancing selection, contrasting with the full-length gene signatures (Fig. 4c, Table 2). On the other side of the spectrum, *Pfs230* and *Pfs48/45* showed strong signs of purifying selection, while *Pfs47* displayed neutrality with moderate levels of purifying selection (Fig. 4c, Table 2). Overall, there was no clear correlation between balancing selection signals with vaccine target type, although a general pattern was observed with the sexual stage antigens mostly showing evidence of purifying selection.

### Protein structures and balancing selection on known epitopes confirm transmission-blocking antigens are not under immune selection

To facilitate the search for effective malaria vaccine targets, we applied three structure-based analyses to the structures of ten vaccine candidate proteins (Table 1): nucleotide diversity of a focal residue’s spatial neighborhood (π*), *Tajima’s D* of a focal residue’s spatial neighborhood (D*), and summarization of experimentally solved antibody-antigen structures (Fig. 5). We then explored if there was a high concentration of sequence diversity and balancing selection signatures within certain domains or patches of each protein. Additionally, we sought to determine if these regions correlated with known epitopes.

**Figure 5.**
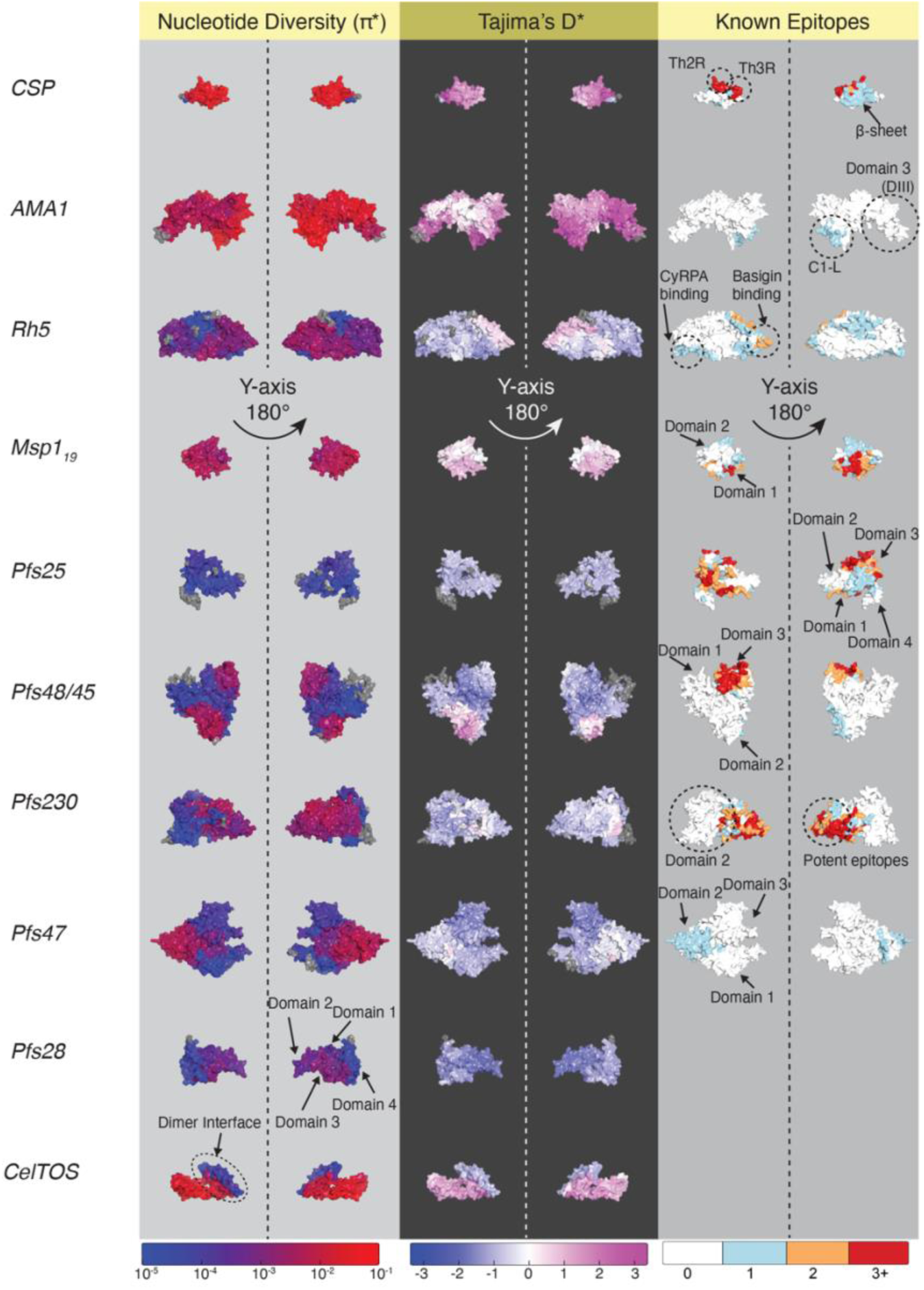
Vaccine candidate structural analyses of nucleotide diversity (π*), Tajima’s D*, and antibody-antigen interactions (epitopes). A 3D sliding window of 15 Å was used for all calculations. Residues with no nearby SNP mapping residues are colored grey. Nucleotide diversity (π*) uses a log scale from extremely low diversity (10^-5^) to extremely high diversity (10^-1^). Tajima’s D* varies from no balancing selection (<0) to strong balancing selection (>2) on a linear scale. In the column of known epitopes, the discrete color scale indicates the number of times a residue was part of an antibody-antigen structure deposited in RCSB. This column is additionally annotated with relevant domain, epitope, and protein-protein interaction surfaces.

The 3D nucleotide diversity, π*, of *CSP, AMA1, CelTOS, and MSP1_19_* showed the overall extreme diversity of the molecules, which agrees with the 2D analysis. The 3D selection measure, *D\**, however, had greater variability across the molecules. *CSP* C-terminal domain exhibited uniformly high diversity and selection, with 95% of spatial windows in regions of π* ≥ 0.01 (maximum π* = 0.046), and 85% and 23% of spatial windows under moderate to strong balancing (D* ≥ 1) and strong balancing selection (D* ≥ 2, maximum D* = 3.34) respectively. The structurally characterized epitopes had similar diversity and selection pressures as the full *CSP* C-terminal domain (Fig. S4). The *D\**, however, could be artificially inflated for parts of the surface, including the conserved β-sheet epitope^74^. Due to the small size of *CSP* C-terminal domain, the 15Å spatial window implemented in biostructmap could include occluded residues on the opposite surface with high polymorphism.

The known epitope of *AMA1*, structurally characterized from antibody 1F9^75^ displayed higher diversity and stronger selection signature than the full *AMA1* structure. Overall, *AMA1* was found to have 58% of spatial windows in regions of π* ≥ 0.01 (maximum π* = 0.058), and 67% were under moderate to strong balancing selection (maximum D* = 3.38), while the epitope of 1F9 shows 100% of their spatial windows in regions of high diversity and high selection. Additionally, we recovered the signature of *AMA1* displaying a distinctive side of the surface with heightened selection, and another side with no selection, termed the “silent face” of *AMA1*^76,77^ (Fig. 5).

*CelTOS* showed a distinctive region of low diversity and selection corresponding to its inferred dimerization interface^78^ (Fig. 5). The overall structure of *CelTOS* was found to have 66% of spatial windows with π* ≥ 0.01 (maximum π* = 0.041), and 55% of spatial windows under moderate to strong balancing selection (maximum D* = 2.01) (Fig. S4).

*MSP1_19_*, *Pfs48/45,* and *Rh5* contained regions under moderate balancing selection (D* between 1-2). *MSP1_19_* domain 1 displayed stronger selection (mean D* = 0.67) than domain 2 (mean D* = 0.34). Among the antibodies with characterized structures, 42D6 is the most potent and targets domains 1 and 2 at the same time^79^. One region in *Rh5* was found to be under selection, which is near its *Basigin* binding interface^80^. Interestingly, this region was shown to be targeted by infection-blocking antibodies^80–82^. For *Pfs48/45*, we found heightened diversity in domains 2 and 3 compared to domain 1, both of which are binding sites of infection-blocking antibodies^83^. Only part of domain 2 was found to be under moderate balancing selection. Interestingly, none of the known epitopes overlap with the selected region.

Among the remaining four vaccine-candidate proteins encompassing the transmission-blocking vaccine class, two had regions under weak balancing selection (0 ≤ D* < 1): *Pfs230* (maximum D* = 0.66) and *Pfs47* (maximum D* = 0.75), and two, *Pfs25* and *Pfs28,* were found to be entirely under purifying selection (D* ≤ 0) with maximum D* of (−0.79) and (−0.78), respectively (Fig. S4).

## Discussion

An ideal malaria vaccine target would be conserved with low balancing selection and containing infection/transmission blocking epitopes that induce strain-transcending antibodies. Against these criteria, ten malaria vaccine antigens targeting sporozoite (n=1), merozoite (n=3) and sexual lifecycle stages (n=6), were assessed against newly generated Zambian parasite genomes, together with publicly available *P. falciparum* WGS spanning five other countries in Africa.

The frequency of unique haplotypes varied across merozoite antigens when considering their full lengths, ranging from a high of 89.3% for *MSP1* to a low of 6.8% for *Rh5*. Notably, transmission-blocking antigens, including *Pfs25*, *Pfs48/45*, *Pfs28*, and *Pfs47*, exhibited markedly lower frequencies of unique haplotypes (ranging from 0.9% to 12.3%) compared to the merozoite antigen *AMA1* (62.5%) and the pre-erythrocytic antigen *CSP* (41.2%). *Pfs230*, another transmission-blocking antigen, demonstrated a frequency of unique haplotypes similar to the highest recorded, *MSP1* (88%). The high number of haplotypes is expected due to the length of the protein (>3100 AAs and 14 domains) and many haplotypes are the result of rare polymorphisms^25^. *CelTOS*, another sexual stage antigen studied, displayed a moderate frequency of unique haplotypes at 33.9%. When focusing on specific regions targeted for vaccine development, the merozoite antigen *AMA1* (Domains I-III) exhibited the highest frequency of unique haplotypes (52.8%). In contrast, other antigens displayed lower frequencies: *Pfs230* (N-terminal prodomain and Domains I-III) at 24.5%, *CSP* (C-terminal) at 17.2%, *Pfs47* (Del2) at 6.1%, *MSP1* (C-terminal region: 19) at 2.8%, and *Pfs48/45* (C-terminal 6-Cys domain) at 2.1%. These findings suggest a lack of a strict pattern based on vaccine types, although transmission-blocking antigens generally exhibited lower frequencies of unique haplotypes compared to other vaccine antigens. Additionally, the observed low frequency unique haplotypes may be attributed to the high malaria transmission intensity in these regions of Africa.

Antibodies with varying effectiveness have been structurally characterized for seven of ten antigens (see Table 1). Our genetic and structural analyses show that the human immune system plays an important role in shaping the haplotype and nucleotide diversity of *P. falciparum* antigens, thus influencing the potency and coverage of the antibodies. Based on the results, we identified five transmission-blocking antigens (*Pfs28*, *Pfs25*, *Pfs48/45*, *Pfs47*, and *Pfs230*) and one blood-stage antigen (*Rh5*) as promising candidates for developing broad-coverage vaccines. *Pfs28* and *Pfs25* demonstrate markedly lower haplotype and nucleotide diversity than other antigens (Table 2; see also ^40,84^) because they are not expressed within human hosts and therefore they are not subjected to immune selection^85,86^. The drawback of no natural exposure to the human immune system from these two antigens is the lack of antibody-boosting effect upon subsequent natural infections. Nevertheless, the low diversity and no signs of selection warrant *Pfs28* and *Pfs25* as promising potential broad-coverage transmission blocking vaccines. Additionally, combining antibodies raised to these antigens results in synergistic transmission blocking activity, highlighting the possibility of a multi-component vaccine using *Pfs25* and *Pfs28*^86^. Other transmission-blocking (sexual stage) antigens, such as *Pfs48/45*, *Pfs47*, and *Pfs230*, also represent promising broad-coverage vaccine candidates, with the added advantage of antibody-boosting upon subsequent exposure, and synergistic transmission blocking activity when combining antibodies of *Pfs48/45* and *Pfs230*^87^. In addition, despite brief exposure to the immune system during merozoite invasion^88^, *Rh5* seems to be a more promising immunogen due to a less diverse haplotype population and potent antibodies with cross-reactivity^80,81^.

For the pre-erythrocytic antigen, *CSP* C-terminal domain was shown to be highly polymorphic in the current study [Fig. 5; see also ^74^]. Notably, the 3D7 haplotype of *CSP* C-terminal domain represents only 3.7% of haplotypes circulating in our study (Fig. 2 and ^29^). Earlier studies have shown that 3D7 haplotype-based RTS,S vaccine efficacy decreases with increased mutational distance from the 3D7 haplotype^33^. *CSP* C-terminal domain has two characterized B-cell epitopes, and both are capable of weak inhibition of hepatocyte invasion^74^. One overlaps with T-cell epitopes Th2R and Th3R and contains many polymorphic residues, which limits the cross-reactivity of antibodies to other *P. falciparum* strains^74,89^. A second epitope on a conserved β-sheet of *CSP* C-terminal domain was shown to have broad cross-reactivity^74^ but much weaker invasion inhibition than antibodies targeting NANP repeats^74,90^. Overall, the C-terminal domain of *CSP* might not be sufficient to elicit full protection for the entire parasite population, namely in generating broad cross-reactive antibodies capable of potent inhibition of infection.

The blood-stage antigens, *AMA1* (Domains I-III) and *MSP1_19_*, are more diverse than the transmission-blocking antigens, with a significant portion of their surface under at least moderate balancing selection (Fig. 5). These results are consistent with previous studies investigating across Africa and globally, with work revealing balancing selection and high haplotype diversity^25,39,91^. Additionally, the 3D7 haplotype was not dominant for these antigens (Fig. 2). Many polymorphic residues in *AMA1* 1F9’s epitope found in our study were shown to abrogate 1F9 binding (H 200 [E,R,L,V], D 204 [K,N], I 225 [N])^75^, implying that a single vaccine is unlikely to confer protection to multiple divergent haplotypes that exist in natural parasite populations. Not included in the present structural analysis is a highly flexible loop of D2, which has a conserved epitope (targeted by 4G2 monoclonal antibody^92^). Among the antibodies with characterized structures for *MSP1_19_,* 42D6 is reported to be the most potent and targets domain 1 and 2 simultaneously^79^. We found three polymorphic residues in the epitope of 42D6, two of which (Pf3D7 E1671K, L1692F) have been confirmed not to significantly impact binding, while the effect on binding and potency of the third residue (Pf3D7 G1623R) remains uncharacterized and thus unknown. In contrast, *Rh5* is uniquely well-conserved among blood-stage antigens, as previously reported^93^. Most potent antibodies to *Rh5* were found around the binding interface of *Rh5* and human protein *Basigin*^81^. These antibodies often target patches with no polymorphisms in our data with one exception (Pf3D7 C203Y), which has been shown to not affect binding^81^. Other epitope regions near *Rh5*’s interface with *PfCyRPA* were not effective in blocking merozoite invasion^82^. Therefore, targeting the *Rh5-Basigin* interface has the potential as an effective vaccine against merozoite invasion.

For transmission-blocking antigens, *Pfs47* was previously shown to be non-essential for *P. falciparum* life cycle, as mutants lacking this protein were capable of normal fertilization^94^. In contrast, both *Pfs48/45* and *Pfs230* are essential antigens for *P. falciparum* life cycle^95,96^. Potent transmission-blocking antibodies have been characterized for *Pfs48/45*^83,97,98^, *Pfs230 (D1, D2)*^99–101^, and *Pfs47*^102^. As reported above, we only found signals of weak or no balancing selection in these three antigens where potent antibodies are binding. Polymorphic residues on domain 3 of *Pfs48/45* in our dataset were found not to affect binding affinity severely and the antibodies retained nanomolar affinity ranges^97^. In *Pfs230*, some of the potent transmission-blocking antibodies raised from natural exposure bound to regions with no polymorphism in our study^100^, while other potent antibodies raised from human vaccine trials bound to polymorphic regions^101^. Among the polymorphic sites, (Pf3D7 G605S, D714N) were shown to affect binding, while (Pf3D7 E654K, E655V, and A699T) did not. The remaining set of polymorphic residues (Pf3D7 N616K, T656N, V727I) segregate at low frequencies and have not been tested for changes in binding affinities. *Pfs47* also contained polymorphic residues in the epitope of a potent antibody^102^. However, the influence of these polymorphisms has not been characterized. Immunization to both *Pfs28* and *Pfs25* have been shown to raise potent transmission-blocking antibodies^86,103–112^. The impact of polymorphic residues (Pf3D7 L63V, V132I) of *Pfs25* on the binding affinities of the antibodies has not yet been characterized.

While a part of transmission-blocking antigens, *CelTOS* is critical to liver invasion by sporozoites during the pre-erythrocytic stage^78^. The heightened balancing selection pressure and nucleotide diversity of *CelTOS* compared to the other transmission-blocking target antigens reflects its intense interaction with the human immune system. While no antibodies have been structurally characterized against *CelTOS* at the time of this study, it has been shown that mice immunized with *CelTOS* can raise potent transmission-blocking antibodies^113^. However, with nearly the entire surface exposed regions of *CelTOS* under moderate balancing selection, it is unlikely to provide broad-coverage vaccinations.

We acknowledge that this study is not without limitations. Firstly, it was a secondary analysis of a broader study^43^, and we did not intend to do an exhaustive study of all vaccine candidate genes. We chose to incorporate representative genes across the three classes of vaccine candidates with a focus on transmission-blocking antigens that are currently under clinical vaccine development. Identifying potential vaccine targets from the proteome is beyond our study scope. Secondly, our study used samples mainly from sub-Saharan Africa with a particular focus on Zambia where genomic data were lacking. However, we acknowledge the absence of representation from Asia or South America. While we recognize that a study with a globally representative set of samples would be more comprehensive, we believe that this investigation is still useful for understanding vaccine development in the context of Africa and Zambia specifically, a country that remains highly malaria endemic. Lastly, the analyses of full-length and partial genes should not be directly compared as they incorporated varying numbers of samples (the partial genes typically included more samples than the full-length genes). The partial gene analyses for regions of interest and the corresponding 3D structural analyses are more relevant for informing vaccine development.

In conclusion, this study offers valuable insights into selecting *P. falciparum* vaccine antigens with regions under low balancing selection and the presence of infection/transmission-blocking epitopes, which are essential for informing the development of new malaria vaccines. Based on these findings, we recommend the following strategies for vaccine design: 1) a multi-stage vaccine encompassing anti-merozoite (*Rh5*) and transmission-blocking antigens (*Pfs25*, *Pfs28*, *Pfs48/45*, *Pfs230*), and 2) transmission-blocking vaccines targeting combinations of *Pfs25*, *Pfs28*, and *Pfs48/45*. These approaches hold promise for the development of more effective malaria vaccines in the future.

## Data Availability

The raw sequence data are available in the NCBI Sequence Read Archive under BioProject PRJNA932927. Key analysis scripts can be accessed at https://github.com/giocarpi/Pf_capture_div_code.

## Contributors

I.I.C, Q.H. and G.C. conceived and designed the study. I.I.C, B.K.B, S.X., Q.H and G.C. generated and collected data. S.X., I.I.C., B.K.B., Q.H., and G.C. analyzed data and interpret results. J.T., Q.H. and G.C. supervised analysis. M.C.M., B.M., C.M., J.M., J.L.S. W.J.M., and D.J.B. contributed to samples/materials. I.I.C., B.K.B., Q.H., and G.C wrote the manuscript. All authors read, edited, and approved the manuscript for publication.

## Acknowledgements

We would like to thank collaborators at the Zambia National Malaria Elimination Centre, the study participants, and the Southern and Central Africa International Centers of Excellence for Malaria Research (ICEMR). We would also like to extend our gratitude to the communities and researchers of malaria endemic countries that enabled the collection and availability of the *P. falciparum* genomes used in this study and made publicly available through the MalariaGEN *P. falciparum* Community. The authors would also like to thank Abebe Fola for his contributions in extracting genomic DNA from DBS samples.

## Supplementary Material

**Figure S1.**
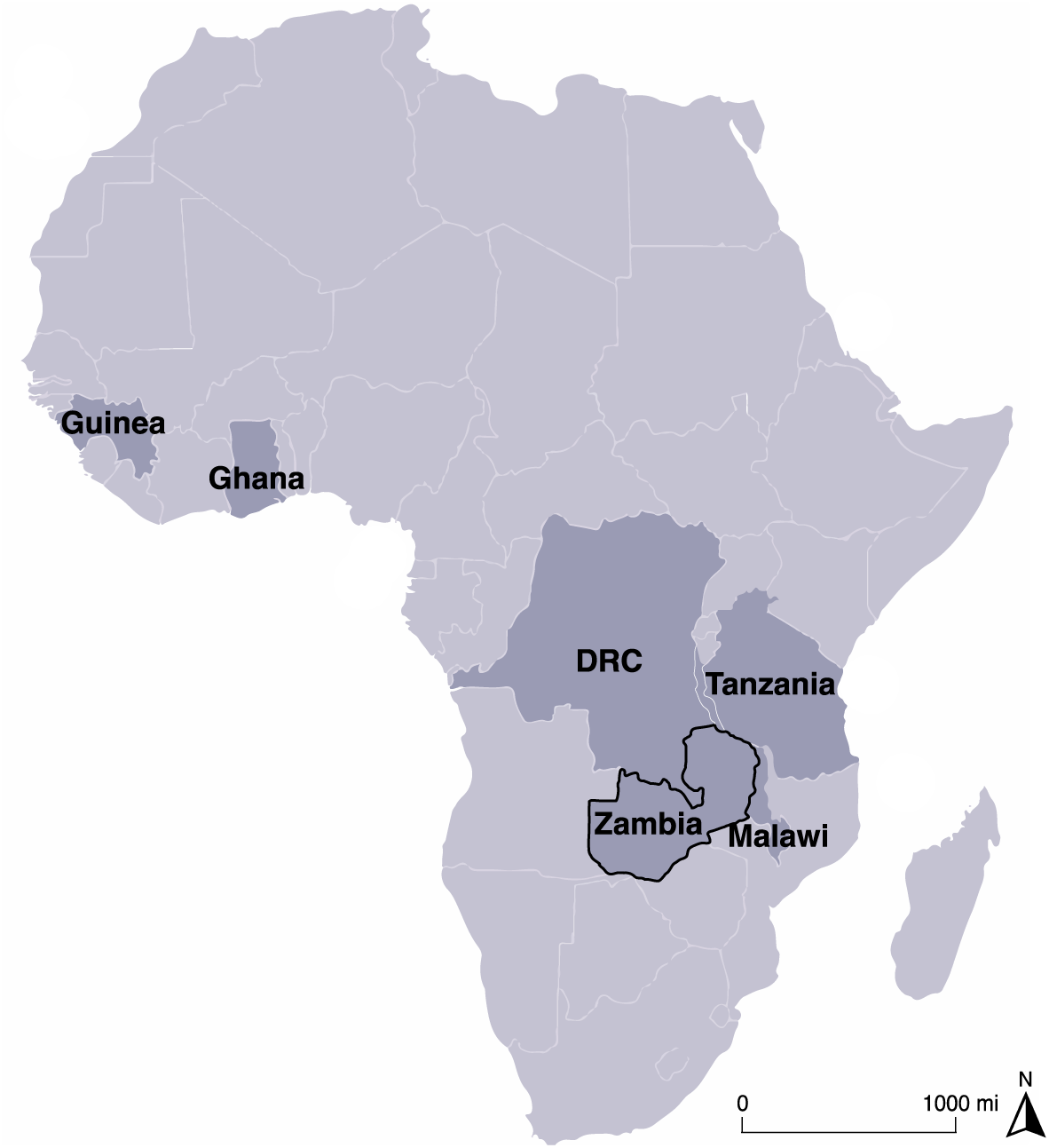
Map of African endemic countries from which *P. falciparum* WGS were included in the study. Map showing the newly generated *P. falciparum* WGS from Zambia (outlined in black) and publicly available WGS data from other malaria endemic countries (MalariaGEN Pf3K) (shown in dark grey).

**Figure S2.**
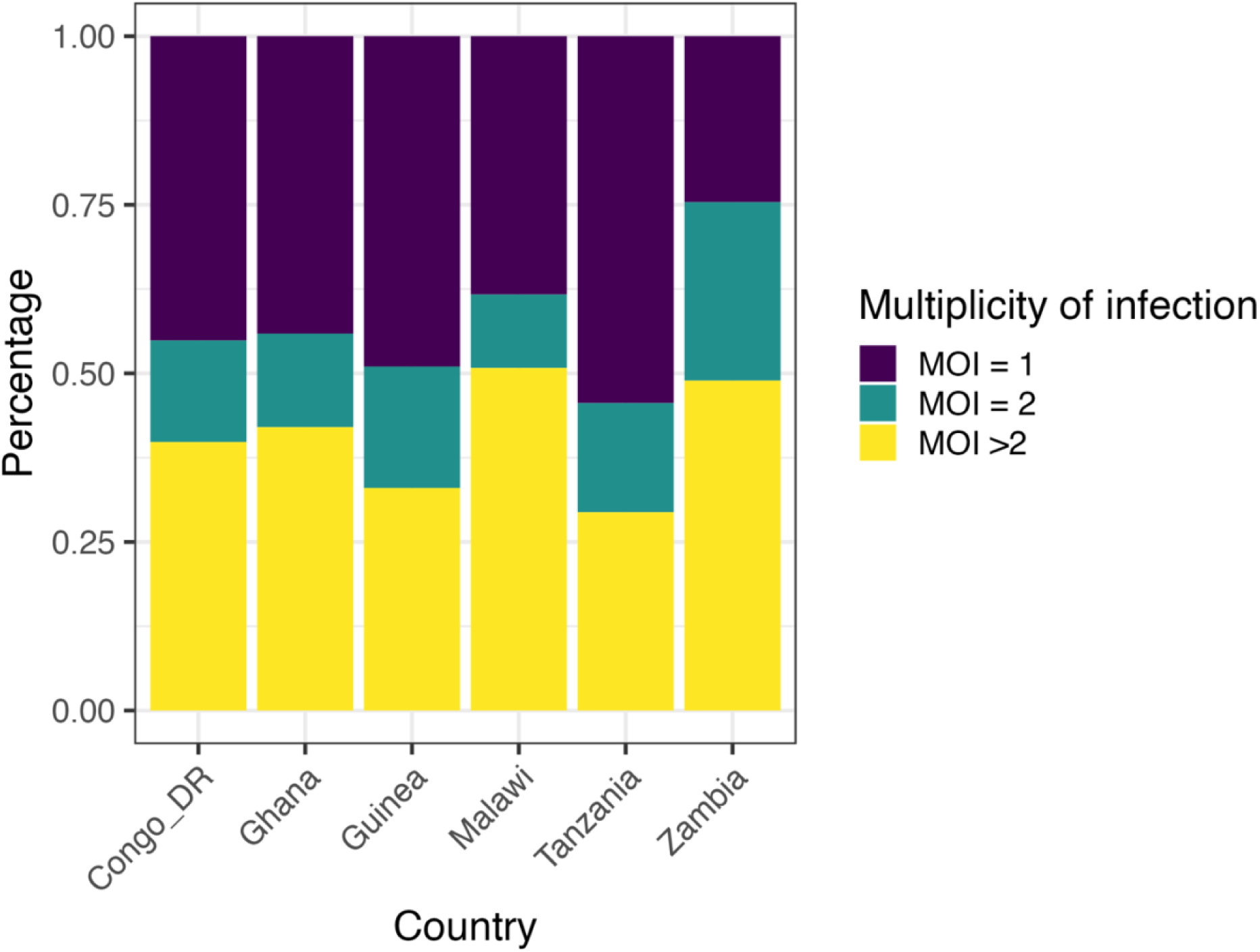
Characterization of multiplicity of infections estimated by *Fws* among 1092 *P. falciparum* genomes across 6 African countries. Colored bars represent the proportion of samples with different levels of multiplicity of infection in each country based on *Fws*. MOI=1: *Fws* above 0.95; MOI=2, *Fws* between 0.8 and 0.95; MOI>2, *Fws* below 0.8.

**Figure S3.**
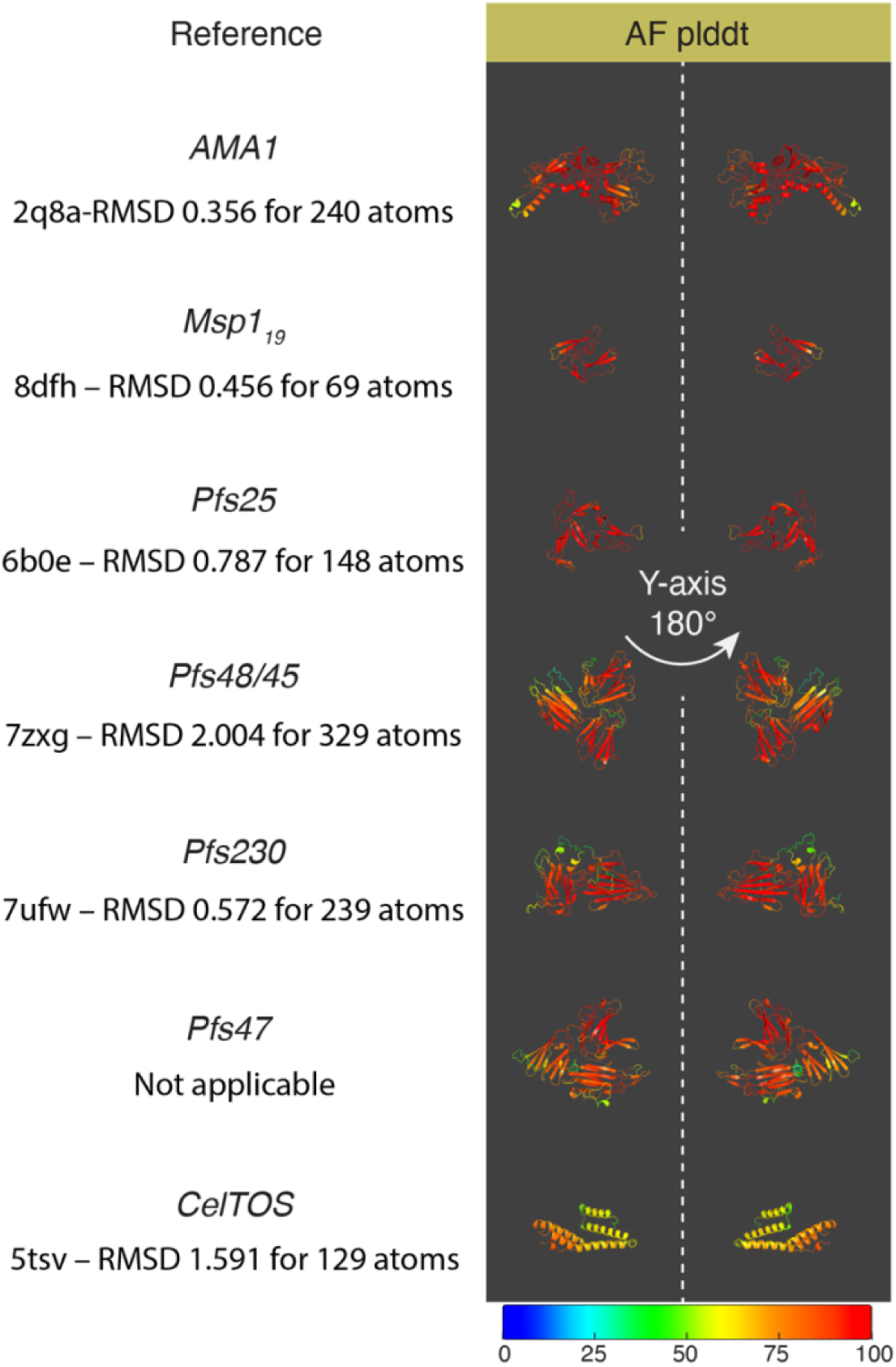
Vaccine candidates’ AlphaFold2 model confidence. The color scale represents AlphaFold2’s reported predicted local Distance Difference Test (pLDDT) for each structure model. See methods for AlphaFold2 implementation. Text under protein names indicates reference structure models and the agreement of alignments measured in Root Mean Squared Deviation (RMSD) over a certain number of atoms.

**Figure S4.**
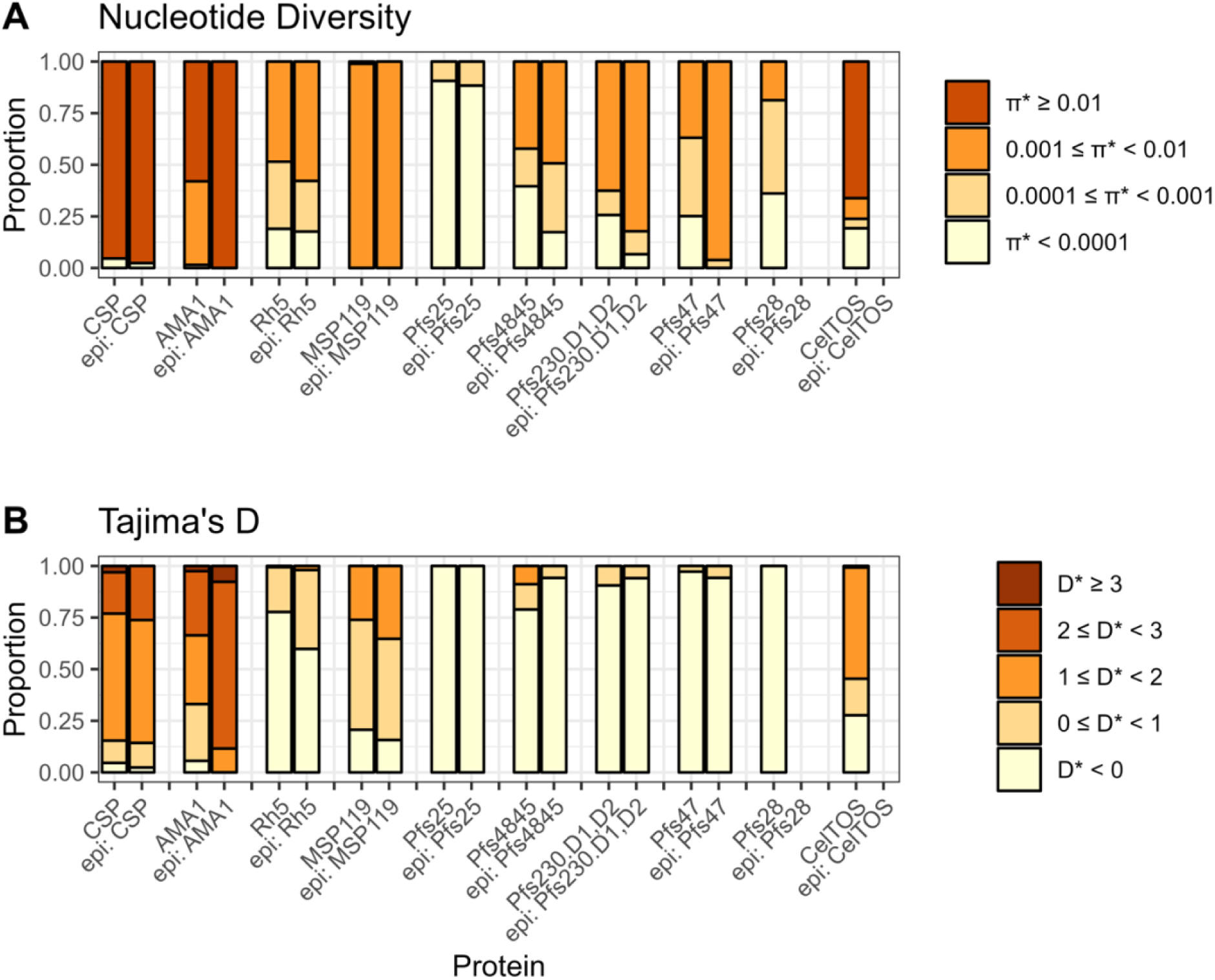
Spatially derived nucleotide diversity and Tajima’s D summaries for whole protein structures versus known epitope residues. Known epitope residues, indicated as “epi: protein name”, are calculated from solved antibody-antigen structures listed in Table 1. Color indicates the proportion of residues in each corresponding bin of nucleotide diversity. See methods for calculation of spatially derived statistics and calculation of epitope residues from solved structures. **A)** Summary of spatially derived nucleotide diversity (π*) values for whole protein structures versus known epitope residues. **B)** Summary of spatially derived Tajima’s D (D*) values for whole protein structures versus known epitope residues.

**Table S1.** Gene information of vaccine candidate antigens. Chromosome number and coordinates, relevant gene symbol and IDs of *P. falciparum* antigens from NCBI gene database.

| Gene Name | Chromosome Number | Chromosome Coordinates | Gene Symbol | Gene ID |
| --- | --- | --- | --- | --- |
| **CSP** | 3 | (221323..222516, complement) | PF3D7_0304600 | 814364 |
| **AMA1** | 11 | (1293856..1295724) | PF3D7_1133400 | 810891 |
| **RH5** | 4 | (1082363..1084150, complement) | PF3D7_0424100 | 812437 |
| **MSP1** | 9 | (1201812..1206974) | PF3D7_0930300 | 813575 |
| **Pfs25** | 10 | (1253417..1254070, complement) | PF3D7_1031000 | 810560 |
| **Pfs45/48** | 13 | (1876016..1877362, complement) | PF3D7_1346700 | 814212 |
| **Pfs230** | 2 | (370438..379845) | PF3D7_0209000 | 812682 |
| **Pfs47** | 13 | (1878875..1880194, complement) | PF3D7_1346800 | 814213 |
| **Pfs28** | 10 | (1251151..1251807, complement) | PF3D7_1030900 | 810459 |
| **CelTOS** | 12 | (659738..660286, complement) | PF3D7_1216600 | 811213 |

